# Mortality in Norway and Sweden before and after the Covid-19 outbreak: a cohort study

**DOI:** 10.1101/2020.11.11.20229708

**Authors:** Frederik E Juul, Henriette C Jodal, Ishita Barua, Erle Refsum, Ørjan Olsvik, Lise M Helsingen, Magnus Løberg, Michael Bretthauer, Mette Kalager, Louise Emilsson

## Abstract

**Objectives:** Norway and Sweden are similar countries regarding ethnicity, socioeconomics and health care. To combat Covid-19, Norway implemented extensive measures such as school closures and lock-downs, while Sweden has been criticised for relaxed measures against Covid-19. We compared the effect of the different national strategies on all-cause and Covid-19 associated mortality.

**Design:** Retrospective cohort.

**Setting:** The countries Norway and Sweden.

**Participants:** All inhabitants.

**Main outcome measures:** We calculated weekly mortality rates (MR) with 95% confidence intervals (CI) per 100,000 individuals as well as mortality rate ratios (MRR) comparing the epidemic year (29^th^ July, 2019 to 26^th^ July, 2020) to the four preceding years (July 2015 to July 2019). We also compared Covid-19 associated deaths and mortality rates for the weeks of the epidemic in Norway and Sweden (16^th^ March to 26^th^ July, 2020).

**Results:** In Norway, mortality rates were stable during the first three 12-month periods of 2015/16; 2016/17 and 2017/18 (MR 14.8 to 15.1 per 100,000), and slightly lower in the two most recent periods including during epidemic period (2018/19 and 2019/20; 14.5 per 100,000). In Sweden, all-cause mortality was stable during the first three 12-month periods of 2015/16; 2016/17 and 2017/18 (MR 17.2 to 17.5 per 100,000), but lower in the year 2018/19 immediately preceding the epidemic (16.2 per 100,000). Covid-19 associated mortality rates were 0.2 per 100,000 (95%CI 0.1 to 0.4) in Norway and 2.9 (95%CI 1.9 to 3.9) in Sweden. The increase in mortality was confined to individuals in 70 years or older.

**Conclusions:** All-cause mortality remained unaltered in Norway. In Sweden, the observed increase in all-cause mortality during Covid-19 was partly due to a lower than expected mortality preceding the epidemic and the observed excess mortality, was followed by a lower than expected mortality after the first Covid-19 wave. This may suggest mortality displacement.

**Strengths and limitations of this study:**

1. Compares two similar contries in all aspects but the handling of the Covid-19 epidemic
2. Evaluates the mortality for several years before and during the epidemic
3. Provides a possible explanation of the observed mortality changes
4. Discusses the socioeconomic effects of the different strategies in the two countries
5. Does not evaluate cause-specific mortality

## Introduction

Covid-19 was declared an international health crisis by the WHO on 30^th^ January, and a pandemic on 11^th^ March, 2020 (1,2). As of 17^th^ September, 945,962 deaths have been officially recorded as a consequence of the disease worldwide (3), and some claim that the death toll is even higher (4,5).

Assessments of the burden of Covid-19 have been hampered by lack of comprehensive data on the disease and of the benefits and harms of the measures against it (6). Cause-specific death rates are prone to bias, especially for a disease with a high asymptomatic burden and large differences in testing and reporting of causes of death between countries. Therefore, all-cause mortality trends may provide a more reliable alternative to assess burden of an epidemic in different countries and regions (4,5).

Norway and Sweden are similar countries regarding ethnicity, governmental and administrative systems, socioeconomics and public health care systems, and both countries have reliable, timely and complete registration of deaths (7,8). To combat the epidemic, Norway implemented extensive measures such as school closures and lock-downs, and reported few Covid-19 associated deaths and little severe disease. Sweden, on the other hand, reported more Covid-19 associated deaths and disease, and has been criticised for their relaxed measures against Covid-19 (9). The substantially different national strategies to control the Covid-19 epidemic in the two countries is a natural experiment that enables difference-in-difference analyses (10) to evaluate the benefits and harms associated with the epidemic and its measures.

To this end, we compared all-cause mortality and Covid-19 associated deaths in Norway and Sweden, before and during the epidemic, in light of the different measures against Covid-19.

## Methods

### Covid-19 and mitigation measures in Norway and Sweden

The first Covid-19 associated death occurred in Norway on 12^th^ March and in Sweden on 11^th^ March, 2020 (11,12). On 12^th^ March, the Norwegian government introduced extraordinary measures against the epidemic (Table 1); emergency laws required closure of all day care centres, schools, universities and other academic institutions, as well as gyms, hair salons, restaurants and movie theatres. Domestic and international travel restrictions, including a total ban for international travel for health care workers, were introduced, and all sport and cultural events as well as all organised sports were cancelled (13,14). The Norwegian government urged the population to stay at home if possible, and contacts with health care services were encouraged only if absolutely necessary (13–16). Most appointments for patients with chronic diseases were cancelled, while some were replaced with telephone and video consultations (17–19).

**Table 1:**
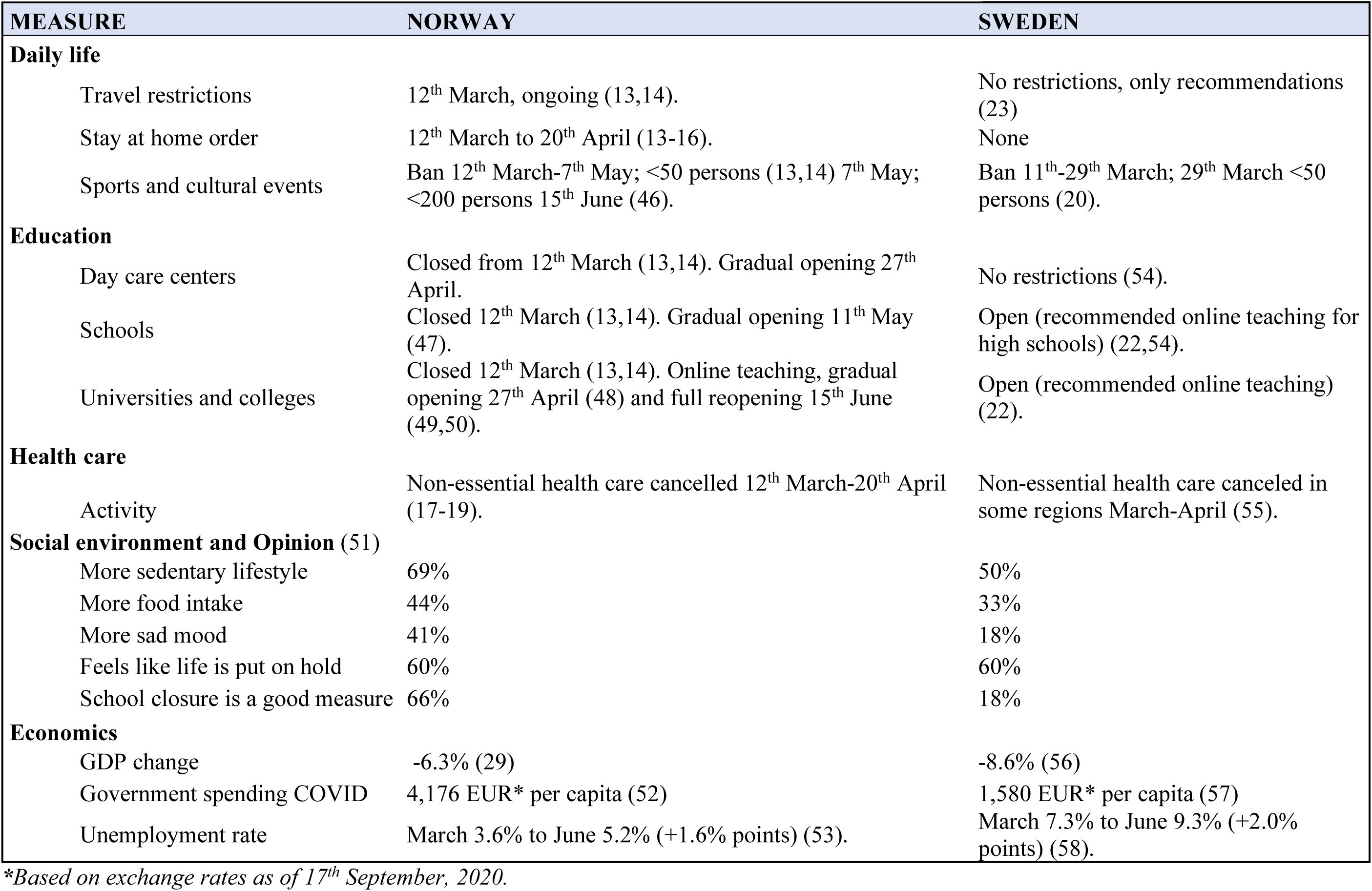
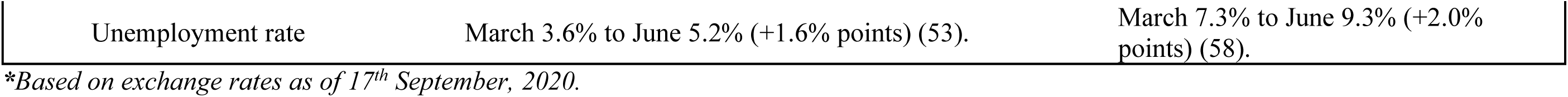
Measures and consequences of Covid-19 in Norway and Sweden.

| MEASURE | NORWAY | SWEDEN |
| --- | --- | --- |
| **Daily life** |  |  |
| Travel restrictions | 12<sup>th</sup> March, ongoing (13,14). | No restrictions, only recommendations (23) |
| Stay at home order | 12<sup>th</sup> March to 20<sup>th</sup> April (13-16). | None |
| Sports and cultural events | Ban 12<sup>th</sup> March-7<sup>th</sup> May; <50 persons (13,14) 7<sup>th</sup> May; <200 persons 15<sup>th</sup> June (46). | Ban 11<sup>th</sup>-29<sup>th</sup> March; 29<sup>th</sup> March <50 persons (20). |
| **Education** |  |  |
| Day care centers | Closed from 12<sup>th</sup> March (13,14). Gradual opening 27<sup>th</sup> April. | No restrictions (54). |
| Schools | Closed 12<sup>th</sup> March (13,14). Gradual opening 11<sup>th</sup> May (47). | Open (recommended online teaching for high schools) (22,54). |
| Universities and colleges | Closed 12<sup>th</sup> March (13,14). Online teaching, gradual opening 27<sup>th</sup> April (48) and full reopening 15<sup>th</sup> June (49,50). | Open (recommended online teaching) (22). |
| **Health care** |  |  |
| Activity | Non-essential health care cancelled 12<sup>th</sup> March-20<sup>th</sup> April (17-19). | Non-essential health care canceled in some regions March-April (55). |
| **Social environment and Opinion (51)** |  |  |
| More sedentary lifestyle | 69% | 50% |
| More food intake | 44% | 33% |
| More sad mood | 41% | 18% |
| Feels like life is put on hold | 60% | 60% |
| School closure is a good measure | 66% | 18% |
| **Economics** |  |  |
| GDP change | -6.3% (29) | -8.6% (56) |
| Government spending COVID | 4,176 EUR* per capita (52) | 1,580 EUR* per capita (57) |

*\*Based on exchange rates as of 17<sup>th</sup> September, 2020.*

In Sweden, measures were considerably less strict (Table 1); on 29^th^ March, the Swedish Government banned public gatherings and events with more than 50 people (20,21). The Public Health Agency recommended high schools and universities to remain open, but teach online if possible (22); non-essential travel should be avoided (23). People with respiratory symptoms should stay at home (24), and some elective surgery was postponed (25), but otherwise the health care service in Sweden operated as before. Restaurants and bars, gyms, hair salons and movie theatres stayed open, and sports and cultural events continued during the epidemic (20,21).

Starting 12^th^ March, both countries strongly emphasised general measures against Covid-19; social distancing with preferred distance of at least 1 meter; hand wash and disinfection, and self-quarantine for all individuals with symptoms suggestive of Covid-19. Contact tracing, isolation and follow-up of Covid-19 transmission was strictly enforced by public health authorities in both countries (26,27).

### Data sources

Norway and Sweden have similar, single-payer, public health care systems with universal coverage. In each country, all residents are assigned an individually unique national registration number, which provides information on sex and date of birth and allows linkage to national registers with data on socioeconomics characteristics, health and disease, hospitalisation and death. Because reporting of death and other health- and socioeconomic variables is mandatory for all residents national registries are close to 100% complete (28). Since its emergence in February 2020, Covid-19 is categorised as a communicable disease with mandatory, immediate reporting of all positive cases to the Norwegian Institute of Public Health in Norway and the Public Health Agency in Sweden.

### Study design

We retrieved weekly numbers of all deaths (regardless of cause) in Norway and Sweden for the last five years until the 26^th^ July, 2020 from Statistics Norway (SSB) (29), the National Board of Health and Welfare (Sweden) (30) and Statistics Sweden (SCB) (31). These registries are complete with three (Norway) and two (Sweden) weeks delay in registration of deaths. 26^th^ July, 2020, was the last observation in the present study.

We calculated mortality for the following five 12-month periods before the endo of the observation period on July 26^th^, 2020. Mondays were defined as the start of each week:

1. 2015/16: 27^th^ July, 2015 to 31^st^ July, 2016;
2. 2016/17: 1^st^ August, 2016 to 30^th^ July, 2017;
3. 2017/18: 31^st^ July, 2017 to 29^th^ July, 2018;
4. 2018/19: 30^th^ July, 2018 to 28^th^ July, 2019;
5. 2019/20: 29^th^ July, 2019 to 26^th^ July, 2020.

The 2019/20 period includes the Covid-19 epidemic and is compared to each preceding 12-month period, and to the mean of the four preceding 12-month periods, for the whole populations of Norway and Sweden. We separately analysed predefined age groups 0-69 years, 70-79 years, and ≥80 years (24). In sensitivity analysis, we explored different weekly cut offs, but the results did not change.

All Covid-19 associated deaths, i.e. deaths among individuals with a positive Covid-19 test up to 30 days before death, until 3^rd^ August, 2020 in both Norway and Sweden, were retrieved from the Institute of Public Health in Norway (32) and the Public Health Agency of Sweden (33), stratified by age. We calculated proportions of Covid-19 associated deaths in each predefined age group.

### Statistics

We calculated weekly MR per 100,000 inhabitants for Norway and Sweden based on the total number of individuals living in Norway on 1^st^ January of the current year (34) and in Sweden on 31^st^ December the previous year (35); e.g. MR for all weeks of 2020 were based on the number of individuals registered on 31^st^ December, 2019 in Sweden, and on 1^st^ January, 2020 in Norway.

We calculated the mean number of deaths and mortality rates per week, and present the lowest and the highest number of deaths throughout the 12-month periods. We compared mortality rates in the 2019/20 period to the four preceding 12-month periods (2015-2019) and calculated mortality rate ratios (MRR). In addition, we calculated all-cause mortality rates and Covid-19 associated mortality rates for the weeks of the epidemic (16^th^ March to 26^th^ July, 2020), using the total number of deaths and all Covid-19 associated deaths, respectively, and the corresponding populations of 2020. We estimated 95% confidence intervals (CI) for MRRs assuming the number of deaths to follow a Poisson distribution.

All data analysed in this report are publicly available, and no ethical approval was needed. Stata 16.1 (StataCorp, College Station, TX, USA) was used for all analyses. We followed the reporting standards set by Strengthening the Reporting of Observational Studies in Epidemiology (STROBE) (Appendix 1) (36).

### Patient and public involvement

No patients nor the public was involved in the design, conduct nor interpretation of this study.

## Results

### Study population

The population in Norway and Sweden increased slightly from 2015 to 2020, from 5,165,802 in Norway and 9,747,355 in Sweden in 2015, to 5,367,580 in Norway and 10,327,589 in Sweden in 2020. The age distribution of the two populations was similar: 88% of the inhabitants of Norway and 85% of the inhabitants of Sweden were 69 years or younger; 8% and 10% of the inhabitants were aged 70-79 years in Norway and Sweden, respectively, and 4% and 5% of the inhabitants were aged 80 years or older, respectively. The mean number of deaths per week varied between 766 (2015/16) and 790 (2016/17) in Norway, and between 1,651 (2018/19) and 1,777 (2019/20) in Sweden (Table 2).

**Table 2:**
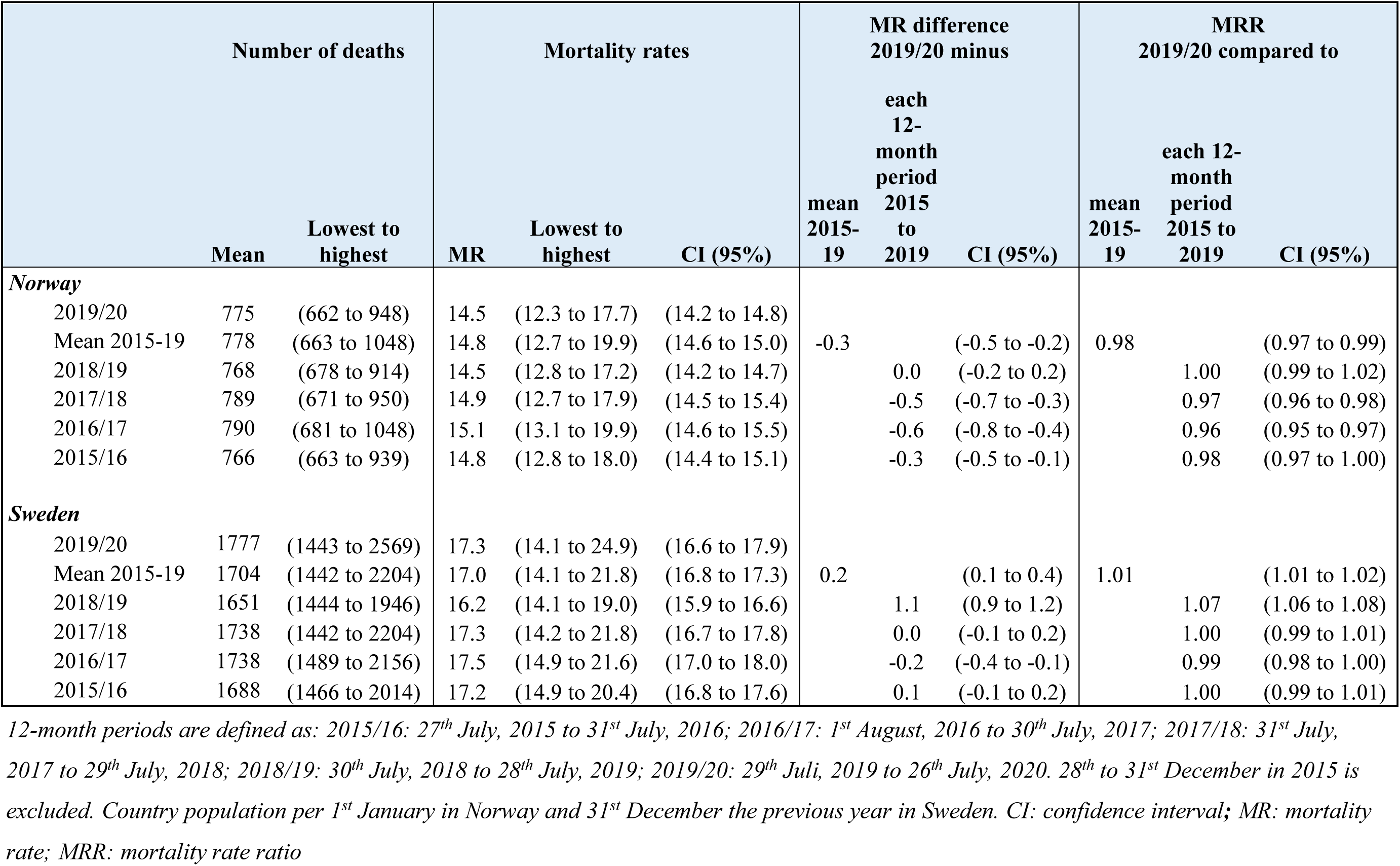
**Weekly number of deaths (mean, lowest and highest weekly numbers), mortality rates (MR) per 100, 000 for 12-month periods, 2015-2020*, and mortality rate ratios (MRR) comparing 2019/20 to the preceding 12-month periods.**

### All-cause mortality

In Norway, mortality rates were stable during the first three 12-month periods (2015/16-2017/18; 14.8 to 15.1 per 100,000), and were slightly lower in the two most recent periods (2018/19 and 2019/20; 14.5 per 100,000) including the epidemic period (Table 2, Figure 1, Figures S1, S2, S3). Similarly, in Sweden, mortality rates were stable during the first three 12-month periods (17.2 to 17.5 per 100,000), and slightly lower in the period preceding the epidemic (16.2 per 100,000). The mortality rate during the year of the epidemic (2019/20) was similar to the first three 12-month periods (MRR 0.99 to 1.00) but was 7% higher than the immediately preceding period (2018/19) (MRR 1.07; 96% CI 1.06 to 1.08).

**Figure 1:**
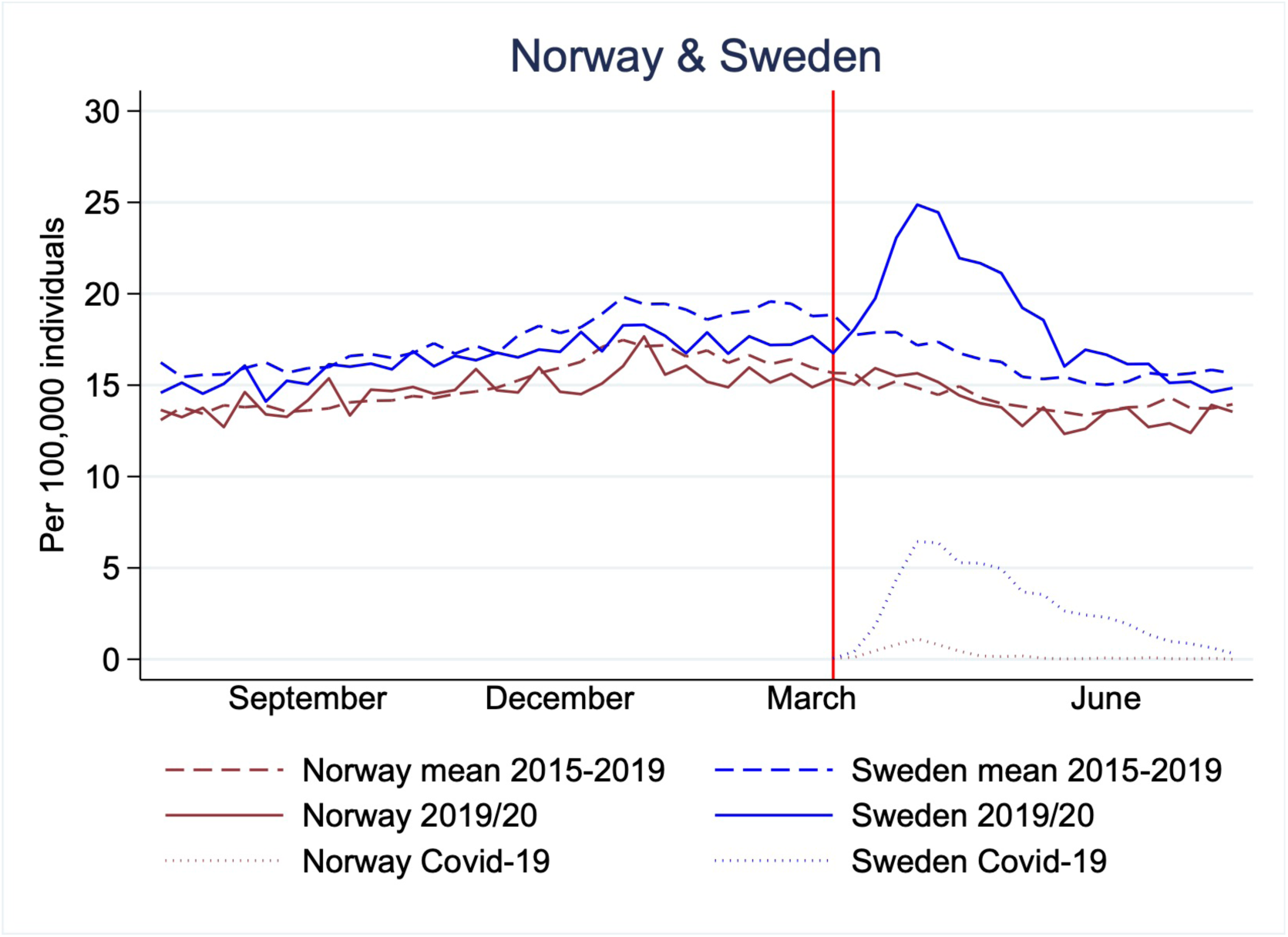
Mortality rates per 100,000 individuals in Norway (brown) and Sweden (blue) for 29^th^ July, 2019 to 26^th^ July, 2020 (solid lines), mean 2015-2019* (dashed lines), and Covid-19 associated mortality rates (dotted). Red vertical line shows the time point for the Covid-19 outbreak in Norway and Sweden (11^th^ and 12^th^ March). Mean is calculated from four 12-month periods defined as: 2015/16: 27^th^ July, 2015 to 31^st^ July, 2016; 2016/17: 1^st^ August, 2016 to 30^th^ July, 2017; 2017/18: 31^st^ July, 2017 to 29^th^ July, 2018; 2018/19: 30^th^ July, 2018 to 28^th^ July, 2019.

In all age groups, the mortality rates during the epidemic period (2019/20) were lower or similar to the mean of the four preceding periods in both countries (Table 3, Figure 2A-C). For individuals 80 and older, mortality rates were highest in 2016/17 in both Norway and in Sweden, while for individuals in Norway younger than 80 years mortality rates were declining from 2015 to 2020 (Table S1, Figures S4, S5). In Sweden, a similar pattern is seen among younger than 70 from 2015-2019.

**Table 3:** **Weekly number of deaths and mortality rates (MR) per age group per 100,000, in 2019/20 and mean of 2015-2019*, and mortality rate ratios (MRR) comparing 2019/20 to the mean 2015 - 2019**.

|  | 2019/20 |  |  |  | Mean 2015-19 |  |  |  | Comparing 2019/20 to mean 2015-19 |  |
| --- | --- | --- | --- | --- | --- | --- | --- | --- | --- | --- |
|  | Number of deaths |  | Mortality rates |  | Number of deaths |  | Mortality rates |  | MRR | CI (95%) |
|  | Mean | Lowest to highest | MR | CI (95%) | Mean | Lowest to highest | MR | CI (95%) |  |  |
| **Norway** |  |  |  |  |  |  |  |  |  |  |
| Age 0-69 | 156 | (126 to 182) | 3.3 | (3.2 to 3.4) | 165 | (123 to 218) | 3.5 | (3.5 to 3.6) | 0.94 | (0.92 to 0.96) |
| Age 70-79 | 177 | (143 to 210) | 41.1 | (40.1 to 42.1) | 165 | (119 to 212) | 43.4 | (42.8 to 44.0) | 0.95 | (0.93 to 0.97) |
| Age ≥ 80 | 442 | (357 to 572) | 194 | (188 to 199) | 449 | (358 to 663) | 202 | (199 to 205) | 0.96 | (0.94 to 0.97) |
| **Sweden** |  |  |  |  |  |  |  |  |  |  |
| Age 0-69 | 285 | (230 to 383) | 3.3 | (3.1 to 3.3) | 301 | (239 to 386) | 3.5 | (3.5 to 3.6) | 0.93 | (0.91 to 0.94) |
| Age 70-79 | 411 | (340 to 585) | 41.7 | (40.4 to 43.5) | 381 | (299 to 491) | 41.7 | (41.2 to 42.2) | 1.00 | (0.99 to 1.02) |
| Age ≥ 80 | 1082 | (830 to 1612) | 204 | (195 to 212) | 1023 | (803 to 1421) | 201 | (198 to 204) | 1.01 | (1.00 to 1.02) |

Mean is calculated from four 12-month periods defined as: 2015/16: 27<sup>th</sup> July, 2015 to 31<sup>st</sup> July, 2016; 2016/17: 1<sup>st</sup> August, 2016 to 30<sup>th</sup> July, 2017; 2017/18: 31<sup>st</sup> July, 2017 to 29<sup>th</sup> July, 2018; 2018/19: 30<sup>th</sup> July, 2018 to 28<sup>th</sup> July, 2019. 28<sup>th</sup> to 31<sup>st</sup> December in 2015 is excluded. Country population per 1<sup>st</sup> January in Norway and 31<sup>st</sup> December the previous year in Sweden. CI: confidence interval; MR: mortality rate; MRR: mortality rate ratio

**Figure 2:**
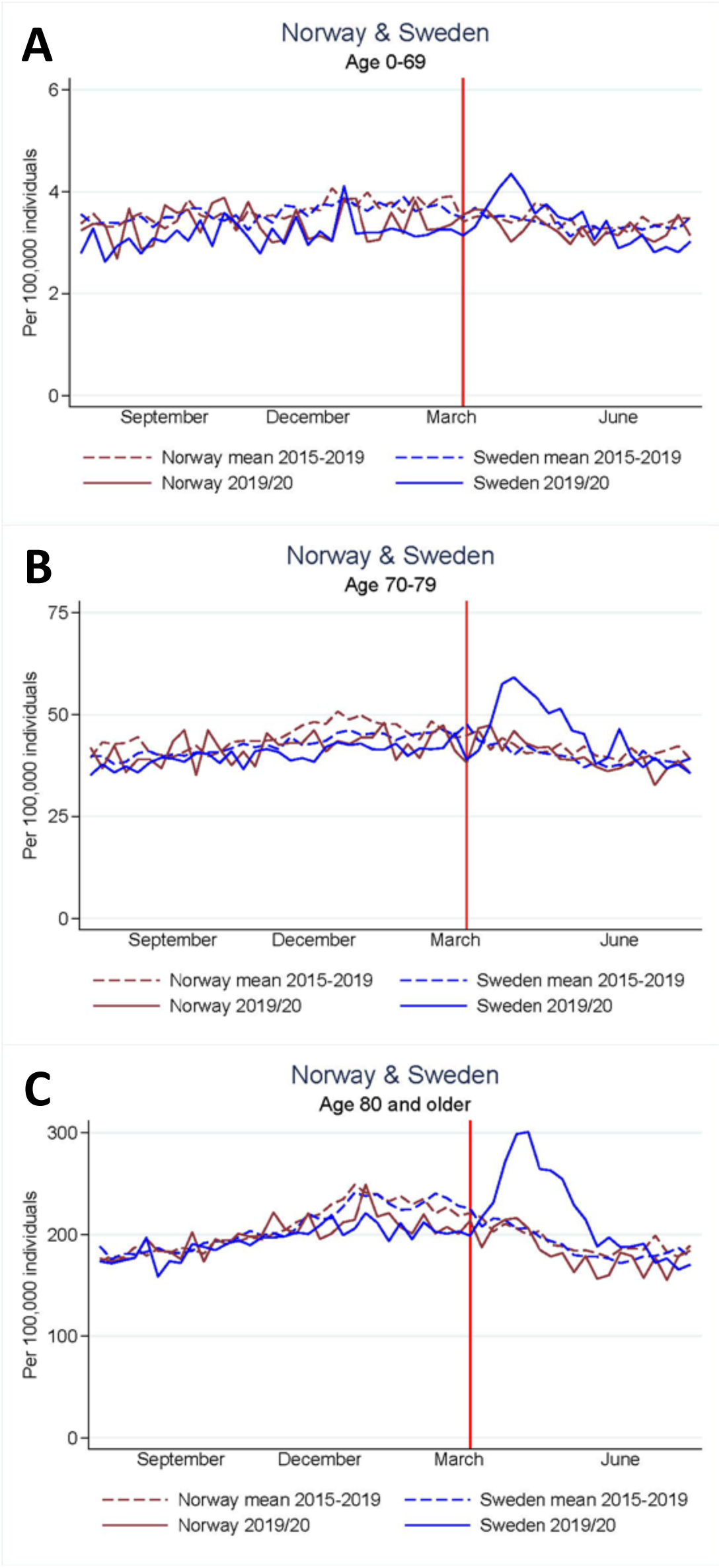
All-cause mortality rates per 100,000 individuals in age groups 0-69 years (A), 70-79 years (B) and ≥ 80 years (C) Norway (brown) and Sweden (blue) for 29^th^ July, 2019 to 26^th^ July, 2020 (solid lines) and mean 2015 to 2019* (dashed lines), and Covid-19 associated mortality rates (dotted). Red vertical line shows the time point for the Covid-19 outbreak in Norway and Sweden (11^th^ and 12^th^ March). Mean is calculated from four 12-month periods defined as: 2015/16: 27^th^ July, 2015 to 31^st^ July, 2016; 2016/17: 1^st^ August, 2016 to 30^th^ July, 2017; 2017/18: 31^st^ July, 2017 to 29^th^ July, 2018; 2018/19: 30^th^ July, 2018 to 28^th^ July, 2019.

### Covid-19 associated mortality

From 11^th^ March to 26^th^ July 2020, a total of 9,125 individuals tested positive for Covid-19, and 255 Covid-19 associated deaths were recorded in Norway, compared to 75,723 Covid-19 positive individuals and 5,741 Covid-19 associated deaths in Sweden (Table S2). The age group distribution of Covid-19 associated deaths was 12.9% (age 0-69), 23.4% (age 70-79) and 63.7% (age 80 and older) in Norway, and 10.9% (age 0-69), 21.5% (age 70-79) and 67.6% (age 80 and older) in Sweden (Table S2). The Covid-19 associated mortality rate was 0.2 per 100,000 individuals (95% CI 0.1 to 0.4) in Norway, and 2.9 per 100,000 individuals (95% CI 1.9 to 3.9) in Sweden (Table S3).

During weeks of 16^th^ March to 26^th^ July 2020, the all-cause mortality rate was 13.9 per 100,000 individuals (95% CI 13.3 to 14.4) in Norway; 0.3 fewer deaths per 100,000 individuals compared to 2015-19. In Sweden, the all-cause mortality rate was 18.7 per 100,000 individuals (95% CI 17.0 to 20.3), 2.3 more deaths per 100,000 individuals compared to 2015-19 (Table S3). This corresponds to 18 fewer deaths in Norway and 232 more deaths in Sweden per week over an 18-week period.

## Discussion

Our study shows that although Covid-19 associated mortality rate was almost 15-fold higher in Sweden than in Norway during the epidemic, all-cause mortality was not higher in Sweden compared with three of the four preceding years. An increase in all-cause mortality was only observed in comparison to the immediately preceding period (2018/19), because mortality was lower than in the previous years. The excess mortality was confined to individuals older than 70 years. In contrast, mortality rates were lower than expected for all ages in Norway and individuals younger than 70 years in Sweden.

At the beginning of the Covid-19 epidemic, extensive social measures were introduced in Norway in the form of restrictions and prohibitions to limit the spread of Covid-19 (13–19), while the public health authorities in Sweden chose a much less intrusive strategy (20–25). The Swedish strategy against Covid-19 has therefore received intense international attention and criticism (9), notably because reported mortality rates in Sweden have been higher than in comparable countries such as Norway. In Sweden, however, mortality was lower than expected in the months preceding the epidemic. This finding may suggest mortality displacement.

Mortality displacement (37) entails temporarily increased mortality (called excess mortality) in a population as a result of external events, such as heat waves (38), or epidemics like influenza (39) or Covid-19. The observed temporary excess mortality likely arises because people in vulnerable groups die weeks or months earlier than they would otherwise, due to the timing and severity of the unusual external event. The excess mortality is therefore preceded or followed by periods of lower than expected mortality. The period preceding the excess mortality in Sweden during the Covid-19 epidemic, characterised by lower mortality than usual, might be due, at least partly, to a mild influenza season during the winter of 2019-20 (40). Further, after the Covid-19 epidemic, we might see a decline in morbidity and mortality below normal levels in Sweden, as the oldest and frailest have already died. Consistent with the theory of mortality displacement and catch-up of the Covid-19 effects in Sweden. Figure 1 indeed indicates a lower than expected mortality rate in Sweden in the last weeks of follow-up in our study. It is too early, however, to reliably estimate this effect.

Mortality rates were lowest in 2019/20 for all ages in Norway. In Sweden 2019/20 was similar to previous year (2018/19) and lower than preceding periods for individuals younger than 70 years. This may be due to successful low-intensity and low-cost measures against Covid-19 such as social distancing, personal hygiene, self-quarantine for individuals with symptoms, and contact tracing in both countries. The rate of registered infectious diseases other than Covid-19 (such as E. coli gastroenteritis and N. gonorrhoea infections) has indeed decreased (41).

Societal burdens related to the measures against Covid-19 were substantially higher in Norway than in Sweden (Table 1). Even though Sweden implemented less restrictive measures than most other countries, the economic ramifications also affected Sweden as part of a global economic system. In the 2^nd^ quarter of 2020 GDP decreased 6.3% in Norway, and 8.6% in Sweden. From March to June 2020, the unemployment rate in Norway increased with 1.6 percentage points from 3.6 to 5.2%, and in Sweden with 2.0 percentage points from 7.3 to 9.3%. Additional government spending, both related to active measures against Covid-19 and mitigation of the economic effects on society, summarize to about 22.4 billion EUR in Norway, and 16.3 billion EUR in Sweden, corresponding to 4,176 EUR per capita in Norway and 1,580 EUR per capita in Sweden, a 2.6-fold difference.

The fight against Covid-19 required extensive health care resources (42,43), and studies from other countries indicate that morbidity and mortality from causes other than Covid-19 have changed after the outbreak of the epidemic (44). A limitation of this study is that we only examined all-cause and Covid-19 associated mortality, and no other specific causes of death, as this information is not yet available for the study period. A Swedish study reviewing the medical records of 122 people (a total of 51% of all deaths in the study-region) found that 70% of the deaths were Covid-19 associated, however only 15% of the cases were indeed Covid-19 specific (45). In addition, the observational study design does not provide an opportunity to draw conclusions about the causal relationship between the social measures in Norway and Sweden and the two countries’ differences in mortality rates.

Our study shows that all-cause mortality was largely unchanged during the epidemic as compared to the previous four years in Norway and Sweden, two countries which employed very different strategies against the epidemic. Excess mortality from Covid-19 may be less pronounced than previously perceived in Sweden, and mortality displacement might explain part of the observed findings. We hope that these findings can pave the way for a less polarized and more open-minded discussion about pros and cons with less compared with more drastic measures against the Covid-19 epidemic.

## Data Availability

All data analysed in this report are publicly available.

## Footnotes

### Author contributions

FEJ, HCJ, IB, ML, MB, MK and LE designed the study, with contribution from all authors. FEJ, HCJ and IB analysed the data. FEJ, HCJ and IB wrote the first draft of the manuscript, and all authors revised the manuscript and approved the final version.

### Transparency declaration

Dr Frederik E Juul affirms that this manuscript is an honest, accurate, and transparent account of the study being reported; that no important aspects of the study have been omitted; and that any discrepancies from the study as planned have been explained.

### License for publication

Dr Frederik E Juul has the right to grant on behalf of all authors and does grant on behalf of all authors, a worldwide licence to the Publishers and its licensees in perpetuity, in all forms, formats and media (whether known now or created in the future), to i) publish, reproduce, distribute, display and store the Contribution, ii) translate the Contribution into other languages, create adaptations, reprints, include within collections and create summaries, extracts and/or, abstracts of the Contribution, iii) create any other derivative work(s) based on the Contribution, iv) to exploit all subsidiary rights in the Contribution, v) the inclusion of electronic links from the Contribution to third party material where-ever it may be located; and, vi) licence any third party to do any or all of the above.

### Ethical approval

Not required.

### Data sharing

No additional data available.

## Appendices

- Appendix 1: STROBE checklist
- Appendix 2: Supplementary tables and figures

## References

1. WHO. Statement on the second meeting of the International Health Regulations (2005) Emergency Committee regarding the outbreak of novel coronavirus (2019-nCoV) [Internet]. [cited 2020 Jun 23]. Available from: https://www.who.int/news-room/detail/30-01-2020-statement-on-the-second-meeting-of-the-international-health-regulations-(2005)-emergency-committee-regarding-the-outbreak-of-novel-coronavirus-(2019-ncov)

2. WHO. WHO Director-General’s opening remarks at the media briefing on COVID-19 - 11 March 2020 [Internet]. [cited 2020 Jun 24]. Available from: https://www.who.int/dg/speeches/detail/who-director-general-s-opening-remarks-at-the-media-briefing-on-covid-1911-march-2020

3. Worldometer. Coronavirus Cases [Internet]. Worldometer. 2020 [cited 2020 Sep 4]. p. 1–22. Available from: https://www.worldometers.info/coronavirus/?

4. Wu J, McCann A, Katz J, Peltier E. 207,000 Missing Deaths: Tracking the True Toll of the Coronavirus Outbreak - The New York Times [Internet]. [cited 2020 Sep 4]. Available from: https://www.nytimes.com/interactive/2020/04/21/world/coronavirus-missing-deaths.html

5. The Economist. Covid-19 data - Tracking covid-19 excess deaths across countries | Graphic detail | The Economist [Internet]. [cited 2020 Sep 4]. Available from: https://www.economist.com/graphic-detail/2020/07/15/tracking-covid-19-excess-deaths-across-countries

6. Vanderweele TJ. Challenges Estimating Total Lives Lost in COVID-19 Decisions: Consideration of Mortality Related to Unemployment, Social Isolation, and Depression [Internet]. Vol. 324, JAMA - Journal of the American Medical Association. American Medical Association; 2020 [cited 2020 Aug 26]. p. 445–6. Available from: https://jamanetwork.com/journals/jama/fullarticle/2768250

7. Pedersen AG, Ellingsen CL. Datakvaliteten i Dødsårsaksregisteret. Tidsskr den Nor Laegeforening [Internet]. 2015 May 5 [cited 2020 Sep 9];135(8):768–70. Available from: https://tidsskriftet.no/en/2015/05/perspectives/data-quality-causes-death-registry

8. Brooke HL, Talbäck M, Hörnblad J, Johansson LA, Ludvigsson JF, Druid H, et al. The Swedish cause of death register. Eur J Epidemiol [Internet]. 2017 Sep 1 [cited 2020 Sep 9];32(9):765–73. Available from: https://pubmed.ncbi.nlm.nih.gov/28983736/

9. BBC News. Did Sweden’s coronavirus strategy succeed or fail? - BBC News [Internet]. [cited 2020 Aug 12]. Available from: https://www.bbc.com/news/world-europe-53498133

10. Kalager M, Zelen M, Langmark F, Adami H-O. Effect of Screening Mammography on Breast-Cancer Mortality in Norway. N Engl J Med [Internet]. 2010 Sep 23 [cited 2020 Sep 9];363(13):1203–10. Available from: http://www.nejm.org/doi/abs/10.1056/NEJMoa1000727

11. Region Stockholm. En person smittad med det nya coronaviruset har avlidit - Region Stockholm [Internet]. [cited 2020 Aug 17]. Available from: https://www.sll.se/verksamhet/halsa-och-vard/nyheter-halsa-och-vard/2020/03/en-person-smittad-med-det-nya-coronaviruset-har-avlidit/

12. NRK. Første koronadødsfall i Norge – NRK Norge – Oversikt over nyheter fra ulike deler av landet [Internet]. [cited 2020 Aug 17]. Available from: https://www.nrk.no/norge/forste-koronadodsfall-i-norge-1.14941788

13. Helsedirektoratet. Helsedirektoratet har vedtatt omfattende tiltak for å hindre spredning av Covid-19 - Helsedirektoratet [Internet]. [cited 2020 Jun 24]. Available from: https://www.helsedirektoratet.no/nyheter/helsedirektoratet-har-vedtatt-omfattende-tiltak-for-a-hindre-spredning-av-covid-19

14. Regjeringen. Omfattende tiltak for å bekjempe koronaviruset [Internet]. regjeringen.no; 2020 [cited 2020 Jun 24]. Available from: https://www.regjeringen.no/no/aktuelt/nye-tiltak/id2693327/

15. FHI. Fastleger og legevakt – smittevern mot covid-19 (koronasykdom) [Internet]. [cited 2020 Jun 30]. Available from: https://www.fhi.no/nettpub/coronavirus/helsepersonell/tiltak-i-primarhelsetjenesten-ved-mistenkt-eller-bekreftet-smitte-med-nytt-/?term=&h=1

16. FHI. Råd til helsepersonell i spesialisthelsetjenesten om covid-19 [Internet]. [cited 2020 Jun 30]. Available from: https://www.fhi.no/nettpub/coronavirus/helsepersonell/tiltak-i-spesialisthelsetjenesten-ved-mistenkt-og-bekreftet-smitte-med-nytt/?term=&h=1

17. Aftenposten. Da Norge stengte: 320.000 færre pasienter fikk sykehushjelp. 24.300 færre operasjoner. Tusenvis av tomme sykehussenger. [Internet]. [cited 2020 Jun 30]. Available from: https://www.aftenposten.no/norge/i/4q9Gqg/da-norge-stengte-320000-faerre-pasienter-fikk-sykehushjelp-24300-faerre-operasjoner-tusenvis-av-tomme-sykehussenger

18. Dagens Medisin. OUS har redusert planlagt aktivitet med over 25 prosent - Nyheter, Spesialisthelsetjeneste, Folkehelse - Dagens Medisin [Internet]. [cited 2020 Jun 30]. Available from: https://www.dagensmedisin.no/artikler/2020/03/31/ous-har-redusert-aktiviteten-med-over-25-prosent/

19. Medisin D. UNN har utsatt 7321 planlagte behandlinger - Nyheter, Spesialisthelsetjeneste - Dagens Medisin [Internet]. [cited 2020 Jun 30]. Available from: https://www.dagensmedisin.no/artikler/2020/03/30/unn-har-utsatt-7321-planlagte-behandlinger/

20. Regeringskansliet. Förordning om förbud mot att hålla allmänna sammankomster och offentliga tillställningar [Internet]. [cited 2020 Aug 11]. Available from: https://www.regeringen.se/artiklar/2020/03/forordning-om-forbud-mot-att-halla-allmanna-sammankomster-och-offentliga-tillstallningar/

21. Polisen. Ytterligare begränsade möjligheter vid sammankomster och tillställningar | Polismyndigheten [Internet]. [cited 2020 Sep 4]. Available from: https://polisen.se/aktuellt/nyheter/2020/mars/ytterligare-begransade-mojligheter-till-allmanna-sammankomster-och-tillstallningar/

22. Folkhälsomyndigheten. Lärosäten och gymnasieskolor uppmanas nu att bedriva distansundervisning — Folkhälsomyndigheten [Internet]. [cited 2020 Aug 11]. Available from: https://www.folkhalsomyndigheten.se/nyheter-och-press/nyhetsarkiv/2020/mars/larosaten-och-gymnasieskolor-uppmanas-nu-att-bedriva-distansundervisning/

23. Folkhälsomyndigheten. Tänk över om resan verkligen är nödvändig — Folkhälsomyndigheten [Internet]. [cited 2020 Aug 11]. Available from: https://www.folkhalsomyndigheten.se/nyheter-och-press/nyhetsarkiv/2020/mars/tank-over-om-resan-verkligen-ar-nodvandig/

24. Folkhälsomyndigheten. Ny fas kräver nya insatser mot covid-19 — Folkhälsomyndigheten [Internet]. [cited 2020 Aug 11]. Available from: https://www.folkhalsomyndigheten.se/nyheter-och-press/nyhetsarkiv/2020/mars/ny-fas-kraver-nya-insatser-mot-covid-19/

25. SVT Nyheter. Nära hälften av alla planerade operationer inställda | SVT Nyheter [Internet]. [cited 2020 Aug 11]. Available from: https://www.svt.se/nyheter/inrikes/nara-halften-av-alla-planerade-operationer-installda

26. Sperre Saunes I, Skau Vegar I, Byrkjeflot H, Lindahl AK, Bråten B. Transition measures: Monitoring and Surveillance [Internet]. [cited 2020 Sep 9]. Available from: https://www.covid19healthsystem.org/countries/norway/livinghit.aspx?Section=1.4Monitoringandsurveillance&Type=Section#29Transitionmeasures:monitoringandsurveillance

27. The Health System Response Monitor (HSRM). Policy responses for Sweden: Monitoring and Surveillance [Internet]. [cited 2020 Sep 9]. Available from: https://www.covid19healthsystem.org/countries/sweden/livinghit.aspx?Section=1.4Monitoringandsurveillance&Type=Section

28. Larsen IK, Småstuen M, Johannesen TB, Langmark F, Parkin DM, Bray F, et al. Data quality at the Cancer Registry of Norway: An overview of comparability, completeness, validity and timeliness. Eur J Cancer. 2009 May 1;45(7):1218–31.

29. YSSB. Norge under Korona/Covid-19 Statistikk om koronakrisen - SSB [Internet]. [cited 2020 Sep 4]. Available from: https://www.ssb.no/korona

30. Socialstyrelsen. Statistik om covid-19 - Socialstyrelsen [Internet]. [cited 2020 Sep 4]. Available from: https://www.socialstyrelsen.se/statistik-och-data/statistik/statistik-om-covid-19/

31. SCB. Corona i statistiken [Internet]. [cited 2020 Sep 4]. Available from: https://www.scb.se/hitta-statistik/corona/corona-i-statistiken/

32. FHI. Ukerapporter om koronavirus og covid-19 - FHI [Internet]. [cited 2020 Aug 11]. Available from: https://www.fhi.no/publ/2020/koronavirus-ukerapporter/

33. Folkhälsomyndigheten. Veckorapporter om covid-19 — Folkhälsomyndigheten [Internet]. [cited 2020 Aug 11]. Available from: https://www.folkhalsomyndigheten.se/folkhalsorapportering-statistik/statistik-a-o/sjukdomsstatistik/covid-19-veckorapporter/

34. SSB. Population - annually, per 1 january - SSB [Internet]. [cited 2020 Sep 9]. Available from: https://www.ssb.no/en/befolkning/statistikker/folkemengde/aar-per-1-januar

35. SCB. Population in the country, counties and municipalities on 31 December 2019 and Population Change in 2019 [Internet]. [cited 2020 Sep 9]. Available from: https://www.scb.se/en/finding-statistics/statistics-by-subject-area/population/population-composition/population-statistics/pong/tables-and-graphs/yearly-statistics--municipalities-counties-and-the-whole-country/population-in-the-country-counties-and-muni

36. Von Elm E, Altman DG, Egger M, Pocock SJ, Gøtzsche PC, Vandenbroucke JP. The Strengthening the Reporting of Observational Studies in Epidemiology (STROBE) Statement: Guidelines for reporting observational studies. UroToday Int J [Internet]. 2009 Oct 18 [cited 2020 Sep 23];2(2):806–8. Available from: www.strobe-statement.org.

37. Huynen MMTE, Martens P, Schram D, Weijenberg MP, Kunst AE. The Impact of Heat Waves and Cold Spells on Mortality Rates in the Dutch Population [Internet]. Vol. 109, Environmental Health Perspectives •. 2001 [cited 2020 Jul 3]. Available from: http://ehpnet1.niehs.nih.gov/docs/2001/109p463-470huynen/abstract.html

38. Kalkstein LS. Direct impacts in cities. Lancet. 1993 Dec 4;342(8884):1397–9.

39. Dushoff J, Plotkin JB, Viboud C, Earn DJD, Simonsen L. Mortality due to influenza in the United States - An annualized regression approach using multiple-cause mortality data. Am J Epidemiol [Internet]. 2006 Jan 15 [cited 2020 Aug 27];163(2):181–7. Available from: https://academic.oup.com/aje/article/163/2/181/95820

40. FHI. Ukerapport Influensa - uke 10, 2020 [Internet]. [cited 2020 Aug 31]. Available from: https://www.fhi.no/contentassets/678386f6617949818dab709c9339090a/vedlegg/2020-10-influensaovervaking-2019-2020-uke-10.pdf

41. Stefanoff P, Løvlie AL, Elstrøm P, Macdonald EA. Reporting of notifiable infectiousdiseases during the COVID-19 response [Internet]. The Journal of the Norwegian Medical Association. 2020 [cited 2020 Sep 23]. Available from: https://tidsskriftet.no/sites/default/files/generated_pdfs/59319-reporting-of-notifiable-infectious-diseases-during-the-covid-19-response.pdf

42. Regjeringen.no. Nasjonale tiltak. 2020 Jul 13 [cited 2020 Aug 11]; Available from: https://www.regjeringen.no/no/tema/Koronasituasjonen/nasjonale-tiltak/id2693684/

43. Helsedirektoratet. Covid-19-samfunnsøkonomisk vurdering av smitteverntiltak-andre rapport Rapport fra ekspertgruppe på oppdrag for Helsedirektoratet.

44. Mafham M, Spata E, Goldacre R, Gair D, Curnow P, Bray M, et al. COVID-19 pandemic and admission rates for and management of acute coronary syndromes in England. Lancet [Internet]. 2020 Aug 8 [cited 2020 Aug 11];396(10248):381–9. Available from: https://www.ctsu.ox.ac.uk/

45. Läkartidningen. Covid-19 oftast inte ensam dödsorsak bland äldre [Internet]. [cited 2020 Sep 1]. Available from: https://lakartidningen.se/aktuellt/nyheter/2020/08/covid-19-oftast-inte-ensam-orsak-vid-dodsfall-bland-aldre/

46. Lovdata. Forskrift om smitteverntiltak mv. ved koronautbruddet (covid-19-forskriften). https://lovdata.no/dokument/SF/forskrift/2020-03-27-470 (accessed Sept 1, 2020).

47. Regjeringen. Skolene åpner for alle elever fra 11. mai. https://www.regjeringen.no/no/aktuelt/skolene-apner-for-alle-elever-fra-11.-mai/id2701512/ (accessed Sept 9, 2020).

48. Regjeringen. Vil åpne samfunnet gradvis og kontrollert. 2020; published online April 7. https://www.regjeringen.no/no/aktuelt/Vil-apne-samfunnet-gradvis-og-kontrollert/id2697060/ (accessed June 24, 2020).

49. Regjeringen. Nå åpner universitetene, høyskolene og fagskolene igjen. https://www.regjeringen.no/no/aktuelt/apning-av-hoyskoler-universitet-og-fagskoler/id2706353/ (accessed Sept 9, 2020).

50. Regjeringen. Spørsmål og svar om koronavirus og studenter i Norge. https://www.regjeringen.no/no/tema/utdanning/innsikt/barnehager-skoler-hoyskoler-og-universiteter-stenges-pa-grunn-av-koronaviruset/koronavirus-og-studenter-i-norge/id2693629/ (accessed Sept 9, 2020).

51. Helsingen LM, Refsum E, Gjostein DK, et al. Trust, threats, and consequences of the COVID-19 pandemic in Norway and Sweden: A comparative survey. medRxiv 2020; : 2020.05.16.20089953.

52. Finansdepartementet. Revidert nasjonalbudsjett 2020 (Meld.St. 2). 2020.

53. SSB. Arbeidskraftundersøkinga, sesongjusterte tal - SSB. https://www.ssb.no/arbeid-og-lonn/statistikker/akumnd (accessed Sept 1, 2020).

54. Folkhälsomyndigheten. Information till förskola, grundskola och gymnasier om covid-19. https://www.folkhalsomyndigheten.se/smittskydd-beredskap/utbrott/aktuella-utbrott/covid-19/verksamheter/information-till-skola-och-forskola-om-den-nya-sjukdomen-covid-19/ (accessed Sept 1, 2020).

55. Schmidt I, Lindblom M, Helmers Barbasso S. Covid-19 har påverkat vårdkontakter, operationer och väntetider. www.socialstyrelsen.se (accessed Sept 9, 2020).

56. SCB. BNP-indikatorn: Kraftig nedgång andra kvartalet 2020. https://www.scb.se/hitta-statistik/statistik-efter-amne/nationalrakenskaper/nationalrakenskaper/nationalrakenskaper-kvartals-och-arsberakningar/pong/statistiknyhet/nationalrakenskaper-2a-kvartalet-2020/ (accessed Sept 1, 2020).

57. The Government of Sweden, Ministry of Finance. Reforms Table 3.1 Measures in additional amending budgets in spring 2020 and proposals in the Spring Amending Budget for 2020 SEK million Measures and proposals 2020 Budget 1.

58. SCB. Arbetskraftsundersökningarna (AKU). https://www.scb.se/hitta-statistik/statistik-efter-amne/arbetsmarknad/arbetskraftsundersokningar/arbetskraftsundersokningarna-aku/ (accessed Sept 1, 2020).

