## Supplementary Appendix for "Mortality in Norway and Sweden before and after the Covid-19 outbreak: a cohort study"

Table of contents:

1. Table S1: Weekly number of deaths, mortality rates and mortality rate ratios per age group for the epidemic year and the previous years
2. Table 2: Covid-19 positive tests and associate deaths in Norway and Sweden
3. Table 3: Weekly number of deaths, mortality rates and mortality rate ratios, for the 18 weeks at the start of the epidemic and the corresponding weeks the years before.
4. Figure S1: All-cause mortality rates per 100,000 individuals in Norway and Sweden for the epidemic year and the previous years
5. Figure S2: Absolute number of deaths in Norway and Sweden for the epidemic year and the average of the previous years
6. Figure S3: Absolute number of deaths in Norway and Sweden for the epidemic year and the previous years
7. Figure S4: Absolute number of deaths in age groups 0-69 years, 70-79 years and ≥ 80 years in Norway and Sweden for the epidemic year and the average of previous years
8. Figure S5: Mortality rates in age groups 0-69 years , 70-79 years and ≥ 80 years in Norway and Sweden for the epidemic year and the average of previous years

*Table S1:* **Weekly number of deaths (mean, lowest and highest weekly numbers), mortality rates (MR) per age group per 100, 000 for 12-month periods, 2015-2020*, and mortality rate ratios (MRR) comparing 2019/20 to the preceding 12-month periods.**

| | | Number of deaths | | Mortality rates | | | MR difference | | MRR | |
|---|---|---|---|---|---|---|---|---|---|---|
| **Age group (years)** | **12-month period** | **Mean** | **Lowest to highest** | **MR** | **Lowest to highest** | **CI (95%)** | **2019/20 minus each 12-month period** | **CI (95%)** | **2019/20 compared to each 12-month period** | **CI (95%)** |
| ***Norway*** | | | | | | | | | | |
| 0-69 | 2019/20 | 156 | (126 to 182) | 3.3 | (2.7 to 3.9) | (3.2 to 3.4) | ref | | ref | |
| 0-69 | 2018/19 | 157 | (131 to 198) | 3.4 | (2.8 to 4.2) | (3.3 to 3.4) | 0.0 | (-0.1 to 0.1) | 0.99 | (0.96 to 1.02) |
| 0-69 | 2017/18 | 164 | (139 to 196) | 3.5 | (3.0 to 4.2) | (3.4 to 3.6) | -0.2 | (-0.3 to -0.1) | 0.95 | (0.92 to 0.98) |
| 0-69 | 2016/17 | 167 | (123 to 215) | 3.6 | (2.7 to 4.6) | (3.5 to 3.7) | -0.3 | (-0.4 to -0.2) | 0.93 | (0.90 to 0.96) |
| 0-69 | 2015/16 | 171 | (138 to 218) | 3.7 | (3.0 to 4.7) | (3.6 to 3.8) | -0.4 | (-0.5 to -0.3) | 0.90 | (0.87 to 0.93) |
| ***Sweden*** | | | | | | | | | | |
| 0-69 | 2019/20 | 285 | (230 to 383) | 3.3 | (2.6 to 4.4) | (3.1 to 3.3) | ref | | ref | |
| 0-69 | 2018/19 | 279 | (239 to 329) | 3.2 | (2.7 to 3.8) | (3.1 to 3.3) | 0.0 | (-0.0 to 0.1) | 1.01 | (0.99 to 1.04) |
| 0-69 | 2017/18 | 298 | (250 to 351) | 3.5 | (2.9 to 4.0) | (3.4 to 3.5) | -0.2 | (-0.3 to -0.1) | 0.94 | (0.92 to 0.96) |
| 0-69 | 2016/17 | 305 | (258 to 348) | 3.6 | (3.1 to 4.1) | (3.5 to 3.6) | -0.3 | (-0.4 to -0.2) | 0.91 | (0.89 to 0.93) |
| 0-69 | 2015/16 | 321 | (268 to 386) | 3.8 | (3.2 to 4.6) | (3.7 to 3.9) | -0.6 | (-0.6 to -0.5) | 0.85 | (0.83 to 0.87) |
| ***Norway*** | | | | | | | | | | |
| 70-79 | 2019/20 | 177 | (143 to 210) | 41.1 | (32.7 to 48.1) | (40.1 to 42.1) | ref | | ref | |
| 70-79 | 2018/19 | 173 | (137 to 209) | 41.9 | (34.1 to 49.8) | (40.9 to 43.0) | -0.8 | (-2.0 to 0.4) | 0.98 | (0.95 to 1.01) |
| 70-79 | 2017/18 | 171 | (130 to 207) | 43.5 | (34.2 to 53.1) | (42.3 to 44.7) | -2.4 | (-3.6 to -1.1) | 0.95 | (0.92 to 0.97) |
| 70-79 | 2016/17 | 163 | (119 to 212) | 44.0 | (33.4 to 56.2) | (42.6 to 45.3) | -2.8 | (-4.1 to -1.6) | 0.94 | (0.91 to 0.96) |
| 70-79 | 2015/16 | 154 | (122 to 199) | 44.3 | (34.3 to 55.9) | (42.9 to 45.6) | -3.1 | (-4.4 to -1.8) | 0.93 | (0.90 to 0.96) |
| ***Sweden*** | | | | | | | | | | |
| 70-79 | 2019/20 | 411 | (340 to 585) | 41.7 | (35.3 to 59.1) | (40.4 to 43.5) | ref | | ref | |
| 70-79 | 2018/19 | 386 | (315 to 439) | 40.1 | (32.2 to 46.6) | (39.3 to 40.9) | 1.6 | (0.8 to 2.4) | 1.04 | (1.02 to 1.06) |
| 70-79 | 2017/18 | 394 | (299 to 491) | 42.8 | (31.9 to 52.4) | (41.6 to 44.0) | -1.1 | (-1.9 to -0.3) | 0.97 | (0.96 to 0.99) |
| 70-79 | 2016/17 | 379 | (320 to 452) | 42.3 | (35.8 to 50.5) | (41.3 to 43.3) | -0.6 | (-1.4 to 0.2) | 0.99 | (0.97 to 1.01) |
| 70-79 | 2015/16 | 365 | (315 to 432) | 41.6 | (35.2 to 48.3) | (40.8 to 42.4) | 0.1 | (-0.7 to 0.9) | 1.00 | (0.98 to 1.02) |
| ***Norway*** | | | | | | | | | | |
| ≥ 80 | 2019/20 | 442 | (357 to 572) | 194 | (155 to 249) | (188 to 199) | ref | | ref | |
| ≥ 80 | 2018/19 | 438 | (363 to 538) | 195 | (163 to 238) | (190 to 200) | -2.0 | (-5.2 to 2.0) | 0.99 | (0.97 to 1.01) |
| ≥ 80 | 2017/18 | 453 | (373 to 576) | 204 | (167 to 259) | (197 to 212) | -11.0 | (-14.6 to -7.4) | 0.95 | (0.93 to 0.96) |
| ≥ 80 | 2016/17 | 461 | (375 to 663) | 209 | (170 to 300) | (200 to 217) | -15.4 | (-19.0 to -11.7) | 0.93 | (0.91 to 0.94) |
| ≥ 80 | 2015/16 | 441 | (358 to 532) | 200 | (162 to 242) | (195 to 206) | -6.9 | (-10.5 to -3.3) | 0.97 | (0.95 to 0.98) |
| ***Sweden*** | | | | | | | | | | |

| | | | | | | | | | | |
|---|---|---|---|---|---|---|---|---|---|---|
| ≥ 80 | 2019/20 | 1082 | (830 to 1612) | 204 | (159 to 301) | (195 to 212) | ref | | ref | |
| ≥ 80 | 2018/19 | 987 | (803 to 1189) | 190 | (154 to 228) | (186 to 195) | 13.5 | (11.2 to 15.9) | 1.07 | (1.06 to 1.08) |
| ≥ 80 | 2017/18 | 1046 | (831 to 1421) | 205 | (162 to 277) | (197 to 213) | 1.2 | (-3.6 to 1.3) | 0.99 | (0.99 to 1.01) |
| ≥ 80 | 2016/17 | 1055 | (869 to 1369) | 209 | (171 to 270) | (202 to 217) | -5.1 | (-7.5 to -2.7) | 0.98 | (0.96 to 0.99) |
| ≥ 80 | 2015/16 | 1003 | (842 to 1242) | 200 | (169 to 248) | (195 to 205) | 3.7 | (1.3 to 6.1) | 1.02 | (1.01 to 1.03) |

**12- month periods are defined as: 2015/16: 27th July, 2015 to 31st July, 2016; 2016/17: 1st August, 2016 to 30th July, 2017; 2017/18: 31st July, 2017 to 29th July, 2018; 2018/19: 30th July, 2018 to 28th July, 2019; 2019/20: 29th Juli, 2019 to 26th July, 2020.*

*28th to 31st December in 2015 is excluded. Country population per 1st January in Norway and 31st December the previous year in Sweden.*
*CI: confidence interval, MR: mortality rate; MRR: mortality rate ratio*

***Table S2:*** **Registered Covid-19 positive tests and associated deaths in Norway and Sweden from 11th March (first Covid-19 associated death) and to 26th July 2020.**

| | | Percentage of total | | |
|---|---|---|---|---|
| | **Total number** | **age 0-69** | **age 70-79** | **≥age 80** |
| ***Norway*** | | | | |
| Covid-19 positive tests | 9,125 | | | |
| Covid-19 associated deaths | 255 | 12.9 | 23.4 | 63.7 |
| ***Sweden*** | | | | |
| Covid-19 positive tests | 75,723 | | | |
| Covid-19 associated deaths | 5,741 | 10.9 | 21.5 | 67.6 |

*Total numbers are per 26th July 2020. Proportions are calculated from cumulated numbers (per age group) at 3rd August 2020.*

***Table S3:*** **Weekly number of deaths and mortality rates (MR) per 100,000, during the epidemic in 2020 (16th March to 26th July) and mean of 18-week periods in 2015-19*, number of registered deaths,[29-31] and mortality rate ratios (MRR) comparing 2020 to the mean 2015-19*.**

| | Number of deaths | | | Mortality rates | | | MR difference | | MRR | |
|---|---|---|---|---|---|---|---|---|---|---|
| | MR | Lowest to highest | CI (95%) | MR | Lowest to highest | CI (95%) | 2020 minus 2015-19 | CI (95%) | 2020 divided by 2015-19 | CI (95%) |
| ***Norway*** | | | | | | | | | | |
| Covid-19 associated | 13.3 | (0 to 60) | (4.8 to 21.7) | 0.2 | (0 to 1.1) | (0.1 to 0.4) | | | | |
| 2020 | 745 | (662 to 855) | (716 to 775) | 13.9 | (12.3 to 15.9) | (13.3 to 14.4) | 0.3 | (-0.6 to -0.1) | 0.98 | (0.96 to 0.99) |
| 2015-19 | 747 | (669 to 905) | (738 to 755) | 14.2 | (12.7 to 17.1) | (14.1 to 14.4) | | | | |
| ***Sweden*** | | | | | | | | | | |
| Covid-19 associated | 302 | (34 to 665) | (201 to 403) | 2.9 | (0.3 to 6.4) | (1.9 to 3.9) | | | | |
| 2020 | 1927 | (1510 to 2569) | (1760 to 2093) | 18.7 | (14.6 to 24.9) | (17.0 to 20.3) | 2.3 | (2.0 to 2.5) | 1.14 | (1.12 to 1.15) |
| 2015-19 | 1639 | (1442 to 1997) | (1614 to 1664) | 16.4 | (14.1 to 19.8) | (16.1 to 16.7) | | | | |

**The periods each year are defined as: 18th March to 28th July, 2019; 19th March to 29th July, 2018; 20th March to 30th July, 2017; 21st March to 30th July, 2016; 16th March to 26th July, 2015.*

*28th to 31st December in 2015 is excluded. Country population per 1st January in Norway and 31st December the previous year in Sweden.*
*CI: confidence interval, MR: mortality rate; MRR: mortality rate ratio*

*Figure S1*: **All-cause mortality rates per 100,000 individuals in Norway (A) and Sweden (B) for 29th July, 2019, to 26th July, 2020 (coloured lines), the corresponding period* the four preceding years (grey lines), and covid-19 associated mortality rates (dotted). Red vertical line shows the time point for the covid-19 outbreak in Norway and Sweden (11th and 12th March).**

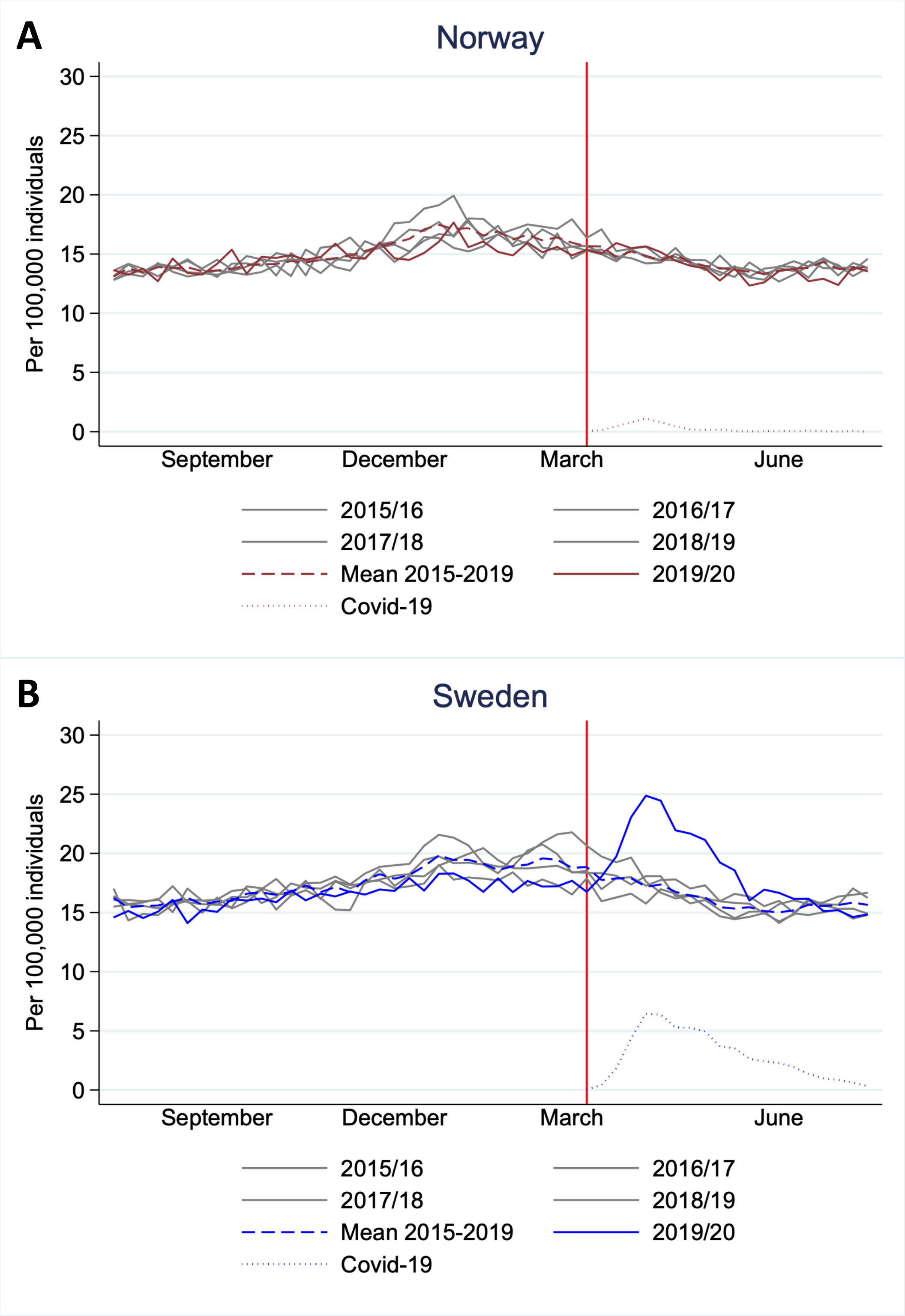

**Previous periods are defined as: 2015/16: 27th July, 2015 to 31st July, 2016; 2016/17: 1st August, 2016 to 30th July, 2017; 2017/18: 31st July, 2017 to 29th July, 2018; 2018/19: 30th July, 2018 to 28th July, 2019.*

***Figure S2:*** **Absolute number of deaths in Norway (brown) and Sweden (blue) for 29th July, 2019 to 26th July, 2020 (solid lines), mean 2015-2019* (dashed lines), and covid-19 associated number of deaths (dotted). Red vertical line shows the time point for the covid-19 outbreak in Norway and Sweden (11th and 12th March).**

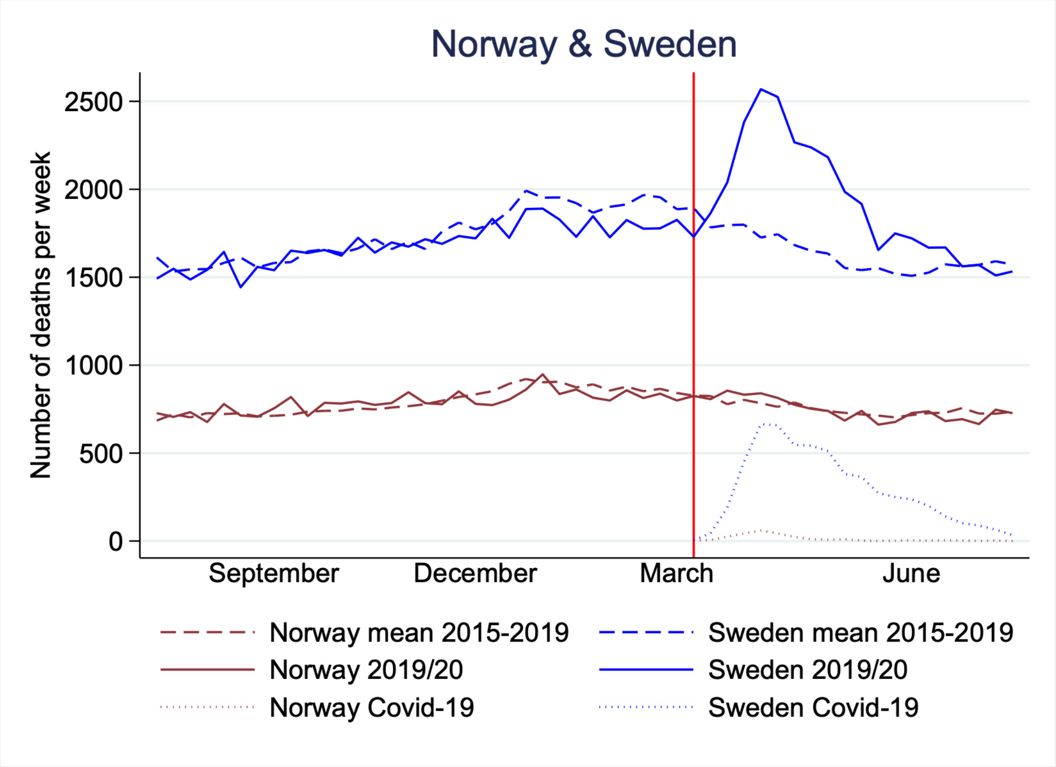

**Mean is calculated from four 12-month periods defined as: 2015/16: 27th July, 2015 to 31st July, 2016; 2016/17: 1st August, 2016 to 30th July, 2017; 2017/18: 31st July, 2017 to 29th July, 2018; 2018/19: 30th July, 2018 to 28th July, 2019.*

*Figure S3:* **Absolute number of deaths in Norway (A) and Sweden (B) for 29th July, 2019 to 26th July, 2020 (coloured lines), the corresponding period* the four preceding years (grey lines), and covid-19 associated deaths (dotted). Red vertical line shows the time point for the covid-19 outbreak in Norway and Sweden (11th and 12th March).**

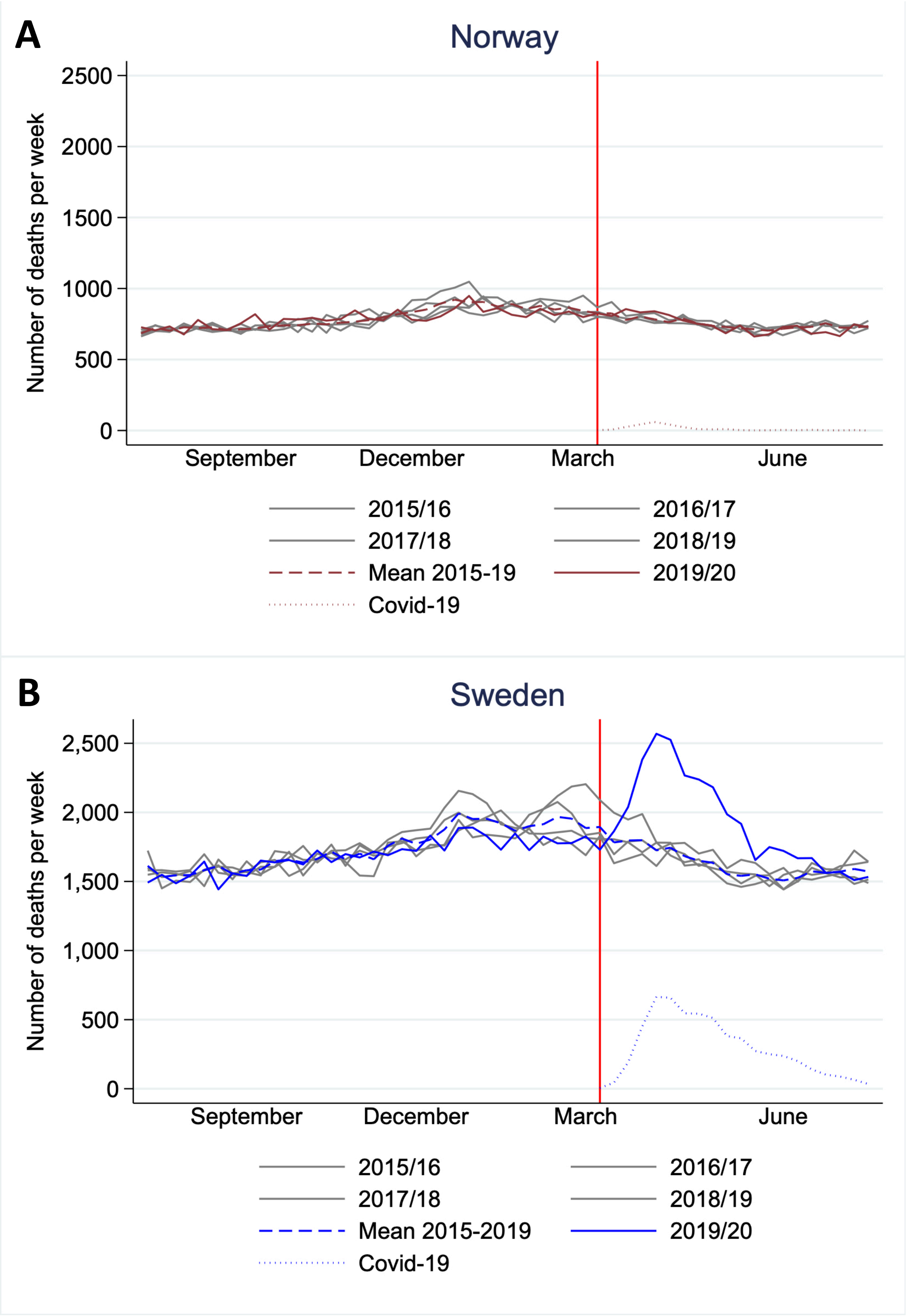

**Previous periods are defined as: 2015/16: 27th July, 2015 to 31st July, 2016; 2016/17: 1st August, 2016 to 30th July, 2017; 2017/18: 31st July, 2017 to 29th July, 2018; 2018/19: 30th July, 2018 to 28th July, 2019.*

***Figure S4:*** **Absolute number of deaths in age groups 0-69 years (A), 70-79 years (B) and ≥ 80 years (C) in Norway (brown) and Sweden (blue) for 29th July, 2019 to 26th July, 2020 (solid lines) and mean 2015-2019* (dashed lines). Red vertical line shows the time point for the covid-19 outbreak in Norway and Sweden (11th and 12th March).**

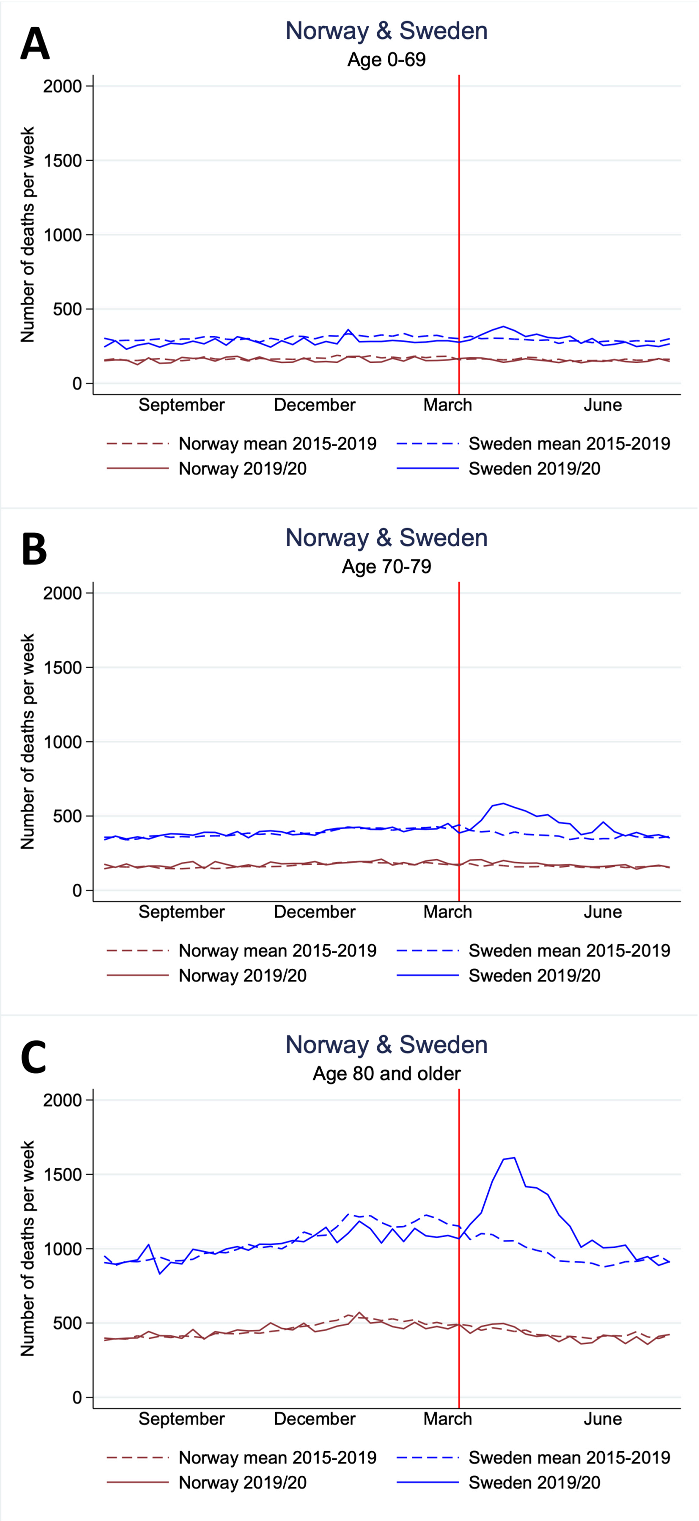

**Mean is calculated from four 12-month periods defined as: 2015/16: 27th July, 2015 to 31st July, 2016; 2016/17: 1st August, 2016 to 30th July, 2017; 2017/18: 31st July, 2017 to 29th July, 2018; 2018/19: 30th July, 2018 to 28th July, 2019*

***Figure S5:*** **Number of deaths in age groups 0-69 years (A), 70-79 years (B) and ≥ 80 years (C) in Norway (brown) and Sweden (blue) for 1st January, 2015 to 26th July, 2020.**

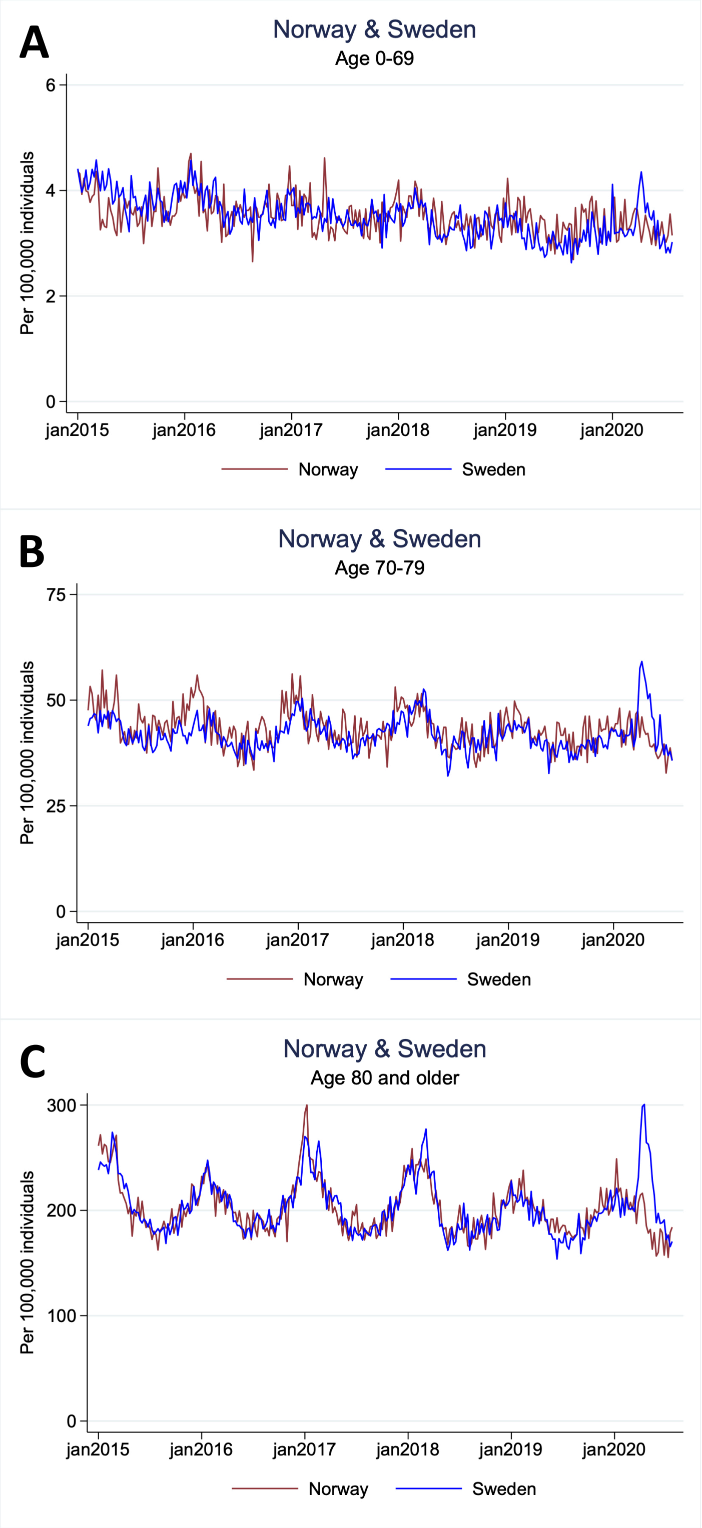
